# Analyzing the Effect of Temperature on the Outspread of COVID-19 around the Globe

**DOI:** 10.1101/2020.05.19.20107433

**Authors:** Pratik Das, Suvendu Manna, Piyali Basak

**Affiliations:** School of Bioscience & Engineering Jadavpur University; School of Engineering, University of Petroleum and Energy Studies

**Keywords:** Covid-19, Environmental effect, Temperature, Total Cases, Recovery, hotspots.

## Abstract

The emergence of the pandemic around the world owing to COVID-19 is putting the world into a big threat. Many factors may be involved in the transmission of this deadly disease but not much-supporting data are available. Till now no proper evidences has been reported supporting that temperature changes can affect COVID-19 transmission. This work aims to correlate the effect of temperature with that of Total Cases, Recovery, Death, and Critical cases all around the globe. All the data were collected in April and the maximum and minimum temperature and the average temperature were collected from January to April (i.e the months during which the disease was spread). Regression was conducted to find a non-linear relationship between Temperate and the cases. It was evident that indeed temperature does have a significant effect on the total cases and recovery rate around the globe. It was also evident from the study that the countries with lower temperatures are the hotspots for COVID-19. The Study depicted a non-linear dose-response between temperature and the transmission, indicating the existence of the best temperature for its transmission. This study can indeed put some light on how temperature can be a significant factor in COVID-19 transmission.

## Introduction

The end of 2019 and the beginning of 2020 witnessed the biggest threat, to the existence of mankind owing to the outspread of the deadly disease COVID-19. The source of this virus is yet a mystery to the whole world(1). A proper vaccine or anti-viral drug is yet to launch in the global market. All this, makes COVID-19 one of the dangerous pandemic, the world has ever faced. Considering the COVID-19 outbreak it is important to recognize its biological physiognomies in the natural environment, especially during its spread. Climate change is an important factor when it comes to the spread of disease(2). Continuous swing in weather conditions and patterns of extreme weather trials is what we refer to climate change. Evidences are there how climate can be a big factor in the spread of disease and how it can have an effect on health(3). Among various environmental factors, temperature could be a vital and distinct factor, which is significant in controlling public health in terms of the epidemic growth and control. Temperature puts forth a diverse influence on people’s living environment in different parts of the world under different climate conditions(4).

SARS-CoV-2 the virus causing COVID-19 is spreading like a fire in the forest. It has affected more than 200 countries in a very short span of 3 months. This deadly disease is having an intense influence on the health care system and economies of affected countries(5). The overall mortality rate is projected to be 6.25 %, but rising to 60% in individuals aged 60 or above. The disease is majorly spread to health care workers, close family members, and individuals near social contacts. As predicted by researchers and doctors the major route of transmission of this deadly disease is through droplets, close direct or indirect contact, but the relative significance of these routes of transmission is presently not clear.

The lack of information can make it hard, to find ways of prevention and control measures. The world saw the first COVID -19 case on December 12, 2019 and soon within 3 months, the World Health Organization(WHO) declared it as a pandemic in March(6). Thus this points out how easily, it is spreading among the community and putting mankind in big suffering. Now this is going to another level of criticalness as it is in the third- and fourth-generation transmission i.e faster human to human transmission. Till now there are not many shreds of evidence, how temperature can be a guiding factor in the virus transmission, or what could be an appropriate range for the same. So a big question which is rumoring all around the world is whether the temperature is at all having any effect on the spread of this deadly disease. There may be some more important or deciding factors for transmission of this virus namely, humidity factor of different countries/places, immunization programs of different countries/government for growing immunity within people of different countries. BCG vaccination may be a probable key factor for resisting the spreading of coronavirus. In India, BCG vaccination is done by the Govt. from a very earlier stage which might play an important factor So far, 11 different types of coronavirus have been identified. Some of them are O, A2, A2A, A3, B, B1 types. The original type was O which was mainly responsible for breaking corona in China. Since 24^th^ January 2020, the corona transmission throughout 60% of the countries of the world are mainly due to the A2A strain. As virulence factor varies greatly from strain to strain, the strain type should be an important variable in statistical analyses. Studies on SARS-CoV showed how transmission of the virus was dependent significantly on the temperature in the city of Beijing and Guangzhou(7) Various research revealed that there was a rise in the daily incidence rate of SARS CoV by 18.18 times when temperature was low compared to that of higher temperatures(8). SARS-CoV-2 virus causing COVID-19 shares structural similarity to that of SARS(9) hence the study of the correlation between Temperature and Transmission could lead us to a significant finding.

This study deals with the correlation of the Total number of Cases, active cases, Deaths, Recovery, and Critical Cases caused globally due to the outspread of COVID -19 with Temperature. We Hypothisized that different temperatures could significantly affect the transmission of the virus. Around 200 countries all around the world have been taken into consideration in this study to find a statistical significance and nonlinear dose-response relationship between the Variable Factors(Total Cases, Deaths, Recovery, and Critical Cases) and temperature. Full sample data was collected for all the affected countries in April and was correlated to their respective average temperature (The Average temperature between Jan to April). Finally, data were analyzed to check whether significant relations exist and if the relatively accurate dose-response relationship could be established.

## Methodology

### Study population

All the confirmed cases all around the world from 204 countries were taken into consideration. The data was collected from the https://www.worldometers.info/coronavirus/ on April 2020 which is now having worldwide data regarding COVID-19(10). Apart from Total Cases (TC) other parameters like Death(D), Recovery (R), and Critical Cases (C) were also taken into consideration[Table S1]. Ln of the total population was considered while analyzing the correlations.

### Average, minimum and maximum temperatures

The minimum temperatures (Min. Temp) and maximum temperatures (Max. Temp) of all the countries in the period of Jan to April were taken into consideration. The average of these two temperatures was considered as average temperature (Avg. Temp). The temperature data was collected from many sources some primary sources are cited as follows (11,12).

### Statistical analysis

A descriptive analysis was performed in Minitab 18.1. Regression was run on the data set to find a polynomial relation between cases and temperature. As mentioned earlier Ln of all the data sets i.e Total Cases (TC), Death(D), Recovery (R), and Critical Cases (C) were taken into consideration. Firstly regression was conducted on Ln(TC) and Max. Temp, Min.Temp Avg.Temp. Similarly regression was run over Ln(R) and Avg. Temp., Ln(D), and Avg.Temp. and Ln(C) and Avg. Temp. The relationships between the three types of temperature data and the number of cumulative total confirmed cases [Ln(N)], was calculated respectively to obtain the fitting equation. After that a non-linear regression was performed on Ln(TC) and Average Temperature based on Generalized Linear Model With Log Link. The Lack of Fit was calculated to predict the accuracy of the model. Later the relationships between other factors i.e Ln(D), Ln(C), and LN(R), and the average temperature was calculated to obtain the fitting equation and the factor dependencies. The main aim of this study is to find a non-linear relationship between the dependence of this disease on temperature.

## Results and Discussion

### Total infection with respect to minimum, maximum and average temperature

As we want to understand the relationship between the total infections with temperature variation in different parts of the globe, the minimum, maximum, and average temperatures of different countries were plotted with the total number of COVID-19 infections in the respective countries and non-linear regression was done on the data. The non-linear plot presented with Figure **Figure 1**, **Figure 2**, **and Figure 3** indicating that total COVID-19 infection changes with minimum, maximum, and average temperature respectively. From the scatter plots it is evident that the countries with high average temperature showed lower infection. Also, countries with a minimum temperature below 40 °F showed a steady COVID-19 infection with irrespective of their region and minimum temperature. Countries with maximum temperature (higher than 50 °F) recorded significantly less amount of COVID-19 positive cases. All these observations possibly pointing to the fact that temperature could be a factor in the fast-spreading of COVID-19 viral infection. The *P*-value was found to be less than 0.001 (p<0.001) except for maximum temperature. On the other hand considering the Residual plot all the residues are normally distributed with a considerable significance in versus fit except that of the maximum temperature. The histogram although it is not skewed uniformly indicating the presence of other factors controlling it. Thus, However, it should be mentioned that countries with immediate governmental action, social maturity, and a healthy lifestyle also affect less COVID-19 positive cases. During the data analysis it was noticed that some of the countries with a lack of proper medical facility and relatively less hygienic lifestyle registered less COVID-19 positive cases. Those countries are found to have a higher average temperature above 70 °F. The quadratic equation representing the relation of Total Cases and Average Temperature can be represented as (where Y represents the total case and x represents Avg. Temp.):

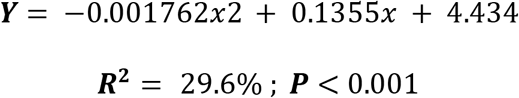

**Figure 1:**
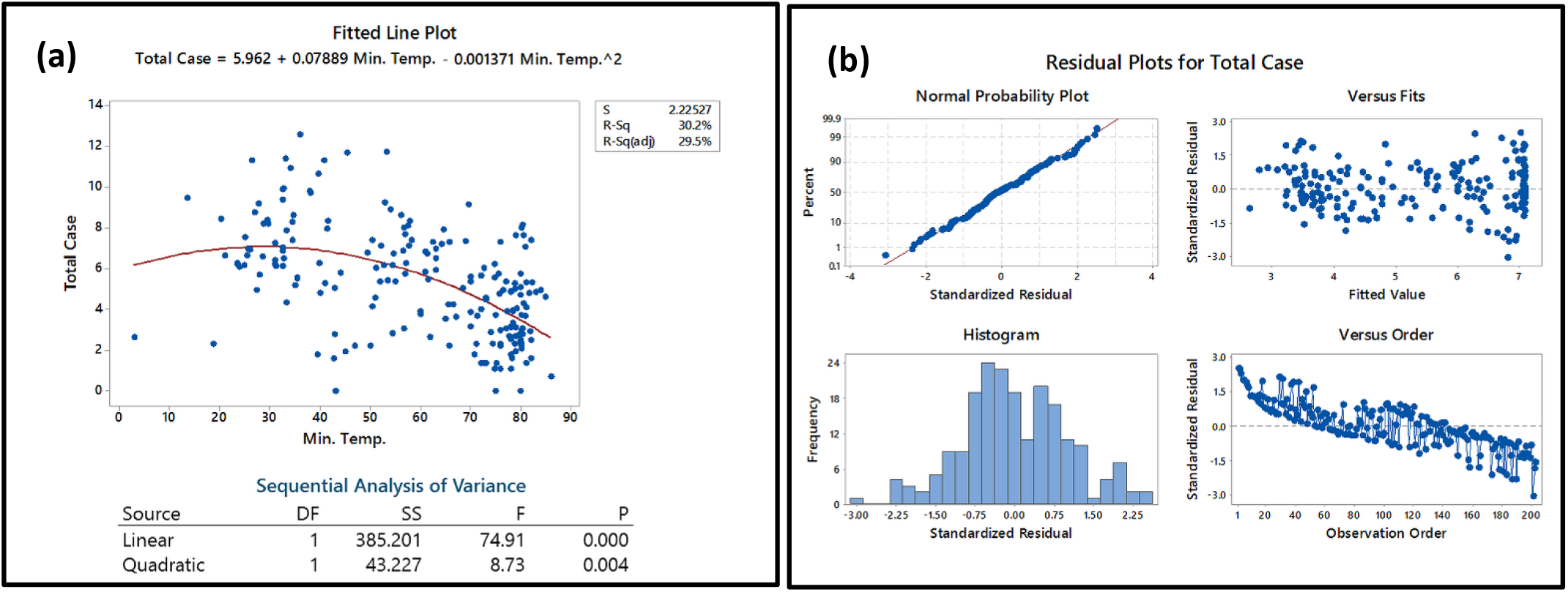
**(a)** The relationship between minimum temperature and Total Cases of COVID-19 transmission in the world; **(b)** The residual plots for the statistical analysis

**Figure 2:**
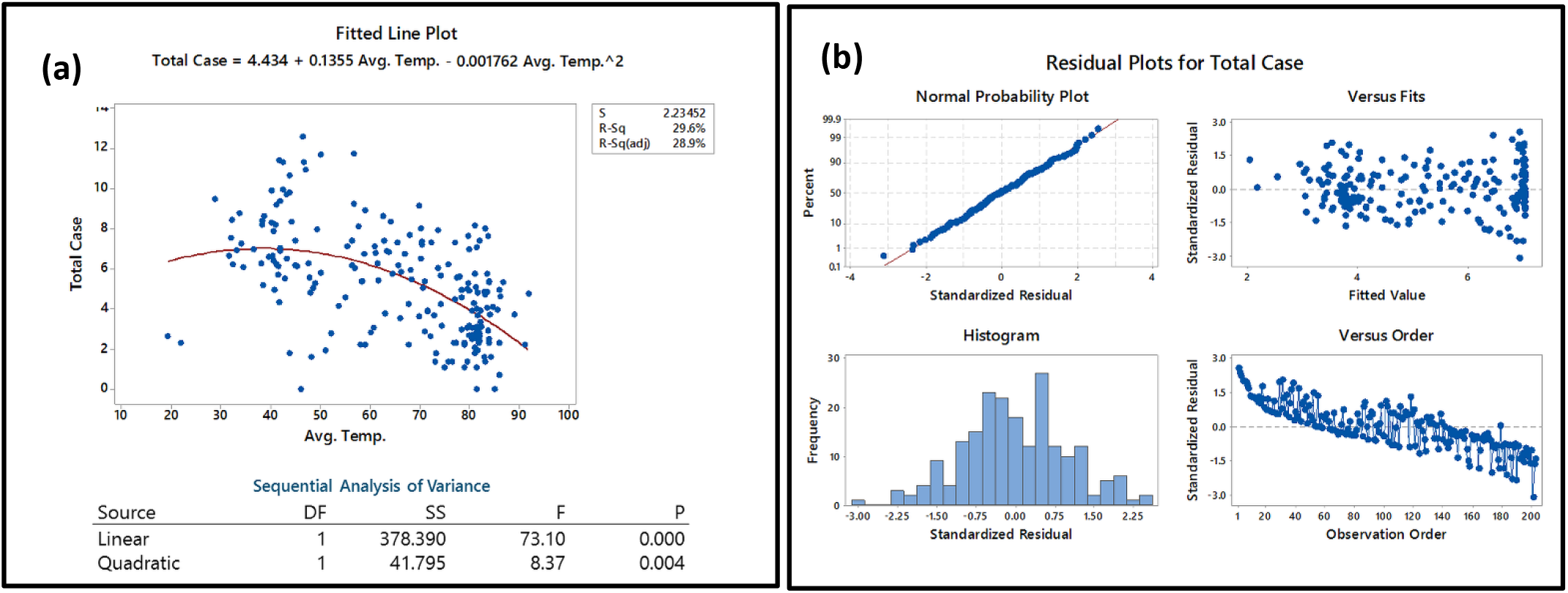
**(a)** The relationship between maximum temperature and Total Cases of COVID-19 transmission in the world **(b)** The residual plots for the statistical analysis

**Figure 3:**
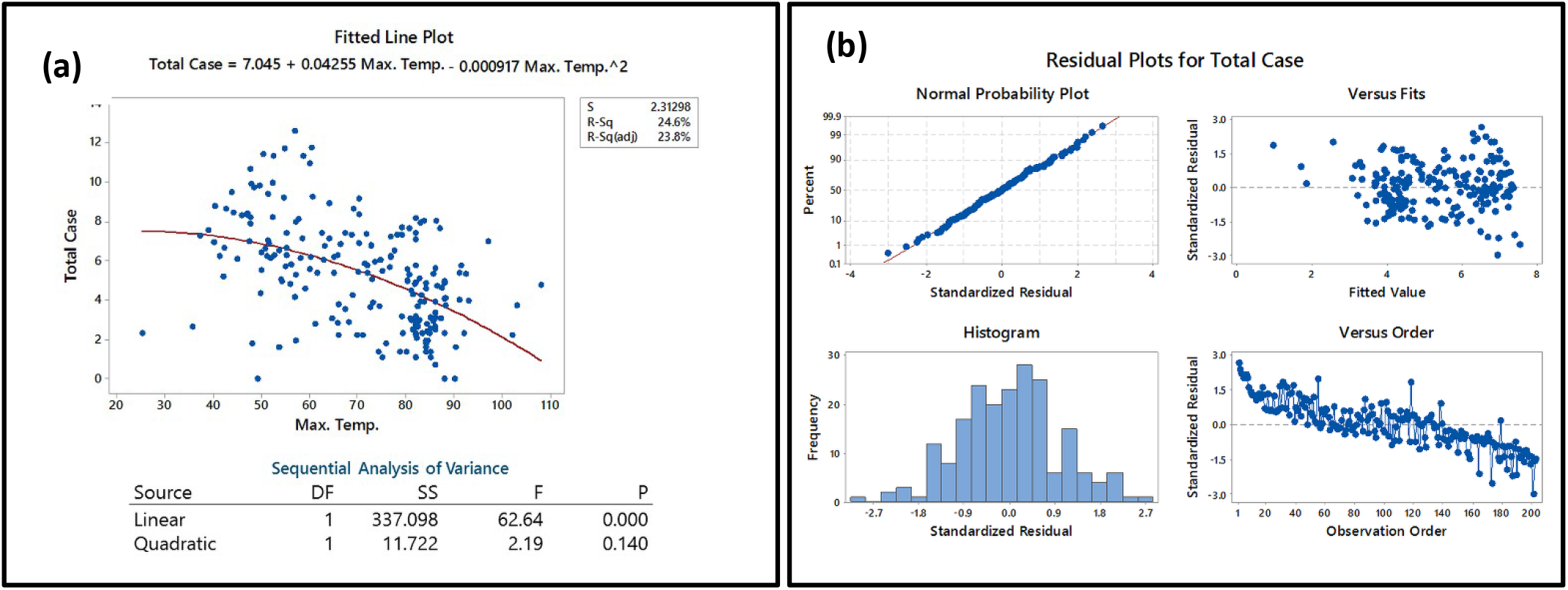
**(a)** The relationship between average temperature and Total Case of COVID-19 transmission in the world **(b)** The residual plots for the statistical analysis

The non-linear regression on Ln(TC) and Average temperature based on Generalized Linear Model With Log Link showed an equational model that can predict the interrelation between the data. The regression fit graph (**Figure 4(a)**) showed an exponential drop in COVID Cases with an increase in temperature. The Lack of Fit test showed the test does not detect any lack-of-fit. The S value of the Model is 2.307, which indicates that the standard deviation of the distance between the data values and the fitted values is approximately 2.307 units. The p-value for the lack-of-fit test is 0.613, which provides no evidence that the model fits the data poorly. The results indicate a highly efficient and probable model for the data. The equation which represents the data is:

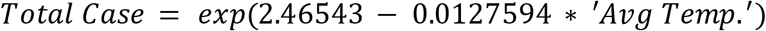

Figure 4(b) represents the residual curve which represents a well-skewed Histogram data which in terms points out the evidential support of the Generalized Linear Model With Log Link model in terms of these data. The normal probability plot here shows a straight line which indicates that the residuals are normally distributed. Residual versus Fit plot shows points fall randomly on both sides of 0, with no recognizable patterns in the points which in term verify the assumption that the residuals are randomly distributed and have constant variance. The residual versus order plot shows points that fall randomly around the centerline with no definite pattern which indicates residues to be independent from one another. Thus overall plot gives a clear result that the model is the best fit for the data.

**Figure 4:**
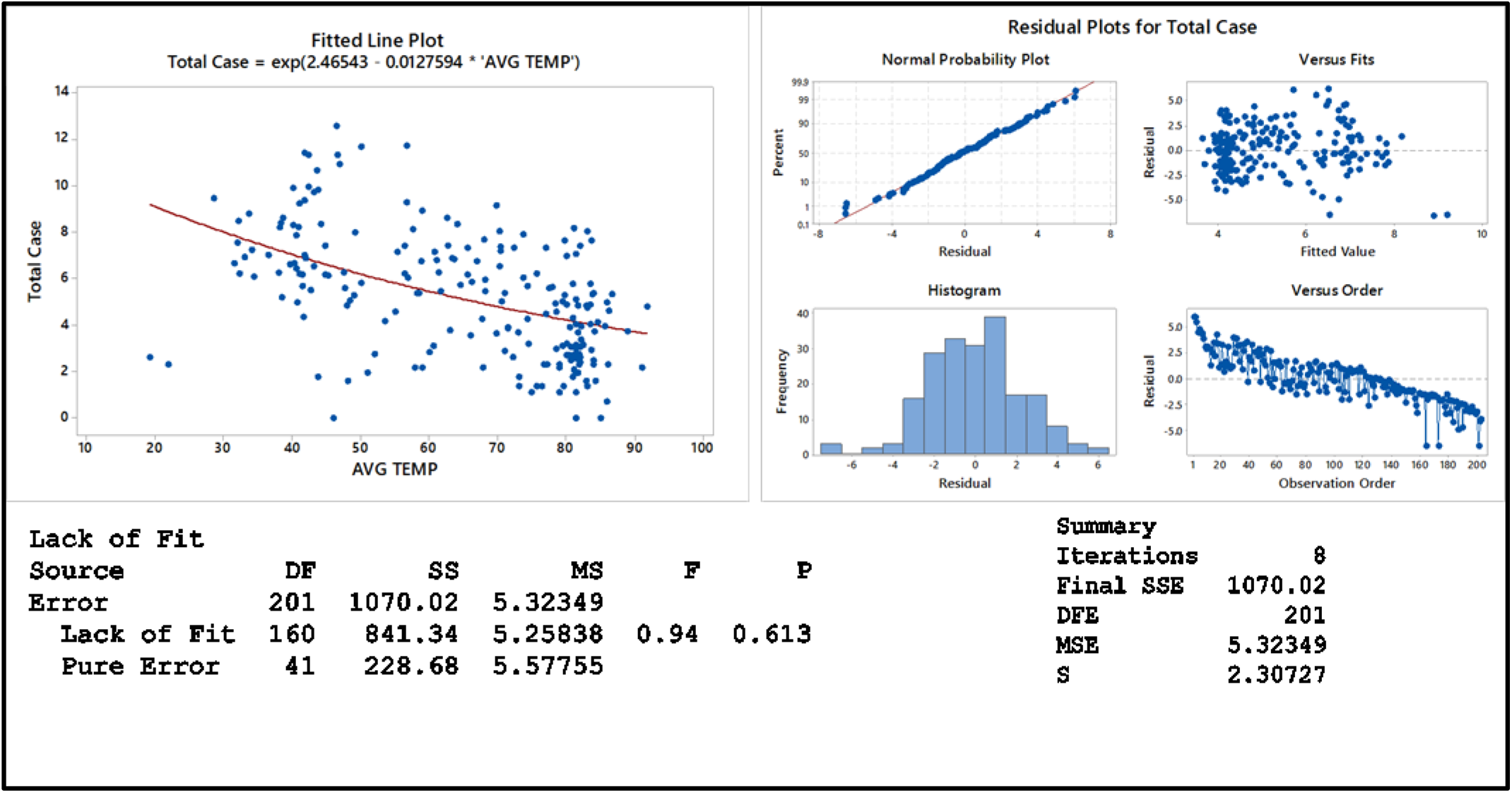
**(a)** The non-linear regression between average temperature and Total Case of COVID-19 transmission in the world **(b)** The residual plots for the statistical analysis

Thus we can conclude that with a 5-degree rise in temperature from---- initially there is an increase in the case but above 40 °F there is a gradual lowering in case numbers. Thus for each 5-degree rise in temp above 40° F there is a lowering in total case by an approx. average of 84 cases per 5 °F. Table 1 gives a detailed calculation. Detailed analysis showed that for each degree rise in temperature above 39 °F (below 40° F) there is a decrease in case number by 19 with a Standard Deviation of ± 14.6[Table S2]. Thus there is a rapid decrease in total cases with temperature increasing above 40° F. [**Figure 5]**. The red dotted line shows the steep decrease in case number with increasing temperature.

**Table1:** Detailed analysis for Temperature and Total case relation

| Temperature[X] | Ln(TC) [Y] | Exp{Ln(TC)}[Total Case] | Difference |
| --- | --- | --- | --- |
| 0 | 4.434 | 84.26781 | - |
| 5 | 5.06745 | 158.7689 | - |
| 10 | 5.6128 | 273.9101 | - |
| 15 | 6.07005 | 432.7023 | - |
| 20 | 6.4392 | 625.9059 | - |
| 25 | 6.72025 | 829.0247 | - |
| 30 | 6.9132 | 1005.46 | - |
| 35 | 7.01805 | 1116.607 | - |
| 40 | 7.0348 | 1135.468 | - |
| 45 | 6.96345 | 1057.275 | 78.19293 |
| 50 | 6.804 | 901.4459 | 155.829 |
| 55 | 6.55645 | 703.7689 | 197.677 |
| 60 | 6.2208 | 503.1056 | 200.6633 |
| 65 | 5.79705 | 329.3266 | 173.7789 |
| 70 | 5.2852 | 197.3937 | 131.933 |
| 75 | 4.68525 | 108.3374 | 89.0563 |
| 80 | 3.9972 | 54.44549 | 53.89186 |
| 85 | 3.22105 | 25.05441 | 29.39108 |
| 90 | 2.3568 | 10.55711 | 14.4973 |
| 95 | 1.40445 | 4.073286 | 6.483829 |
| 100 | 0.364 | 1.439074 | 2.634212 |

**Figure 5:**
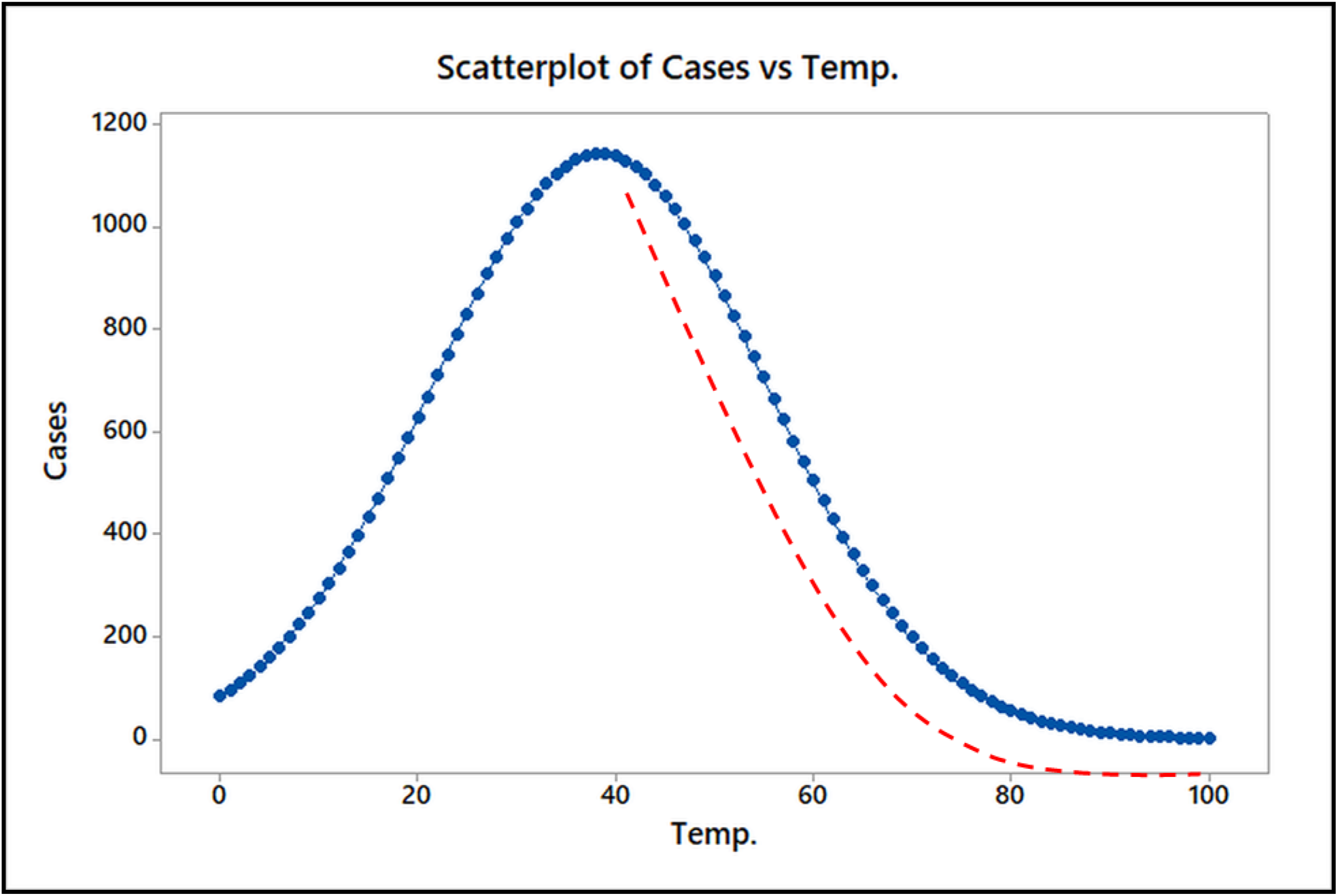
A plot for correlation between the increase in temperature with that of Total Cases A World heat map also depicts how regions with lower temperatures are becoming hotspots for COVID-19 outspread although few exceptions (for Example Brazil) do exist in **Figure 6**. Fast mutation rate and environmental adaptability of the SARS-CoV-2 virus also responsible for uncontrol COVID-19 infections in many countries.

**Figure 6:**
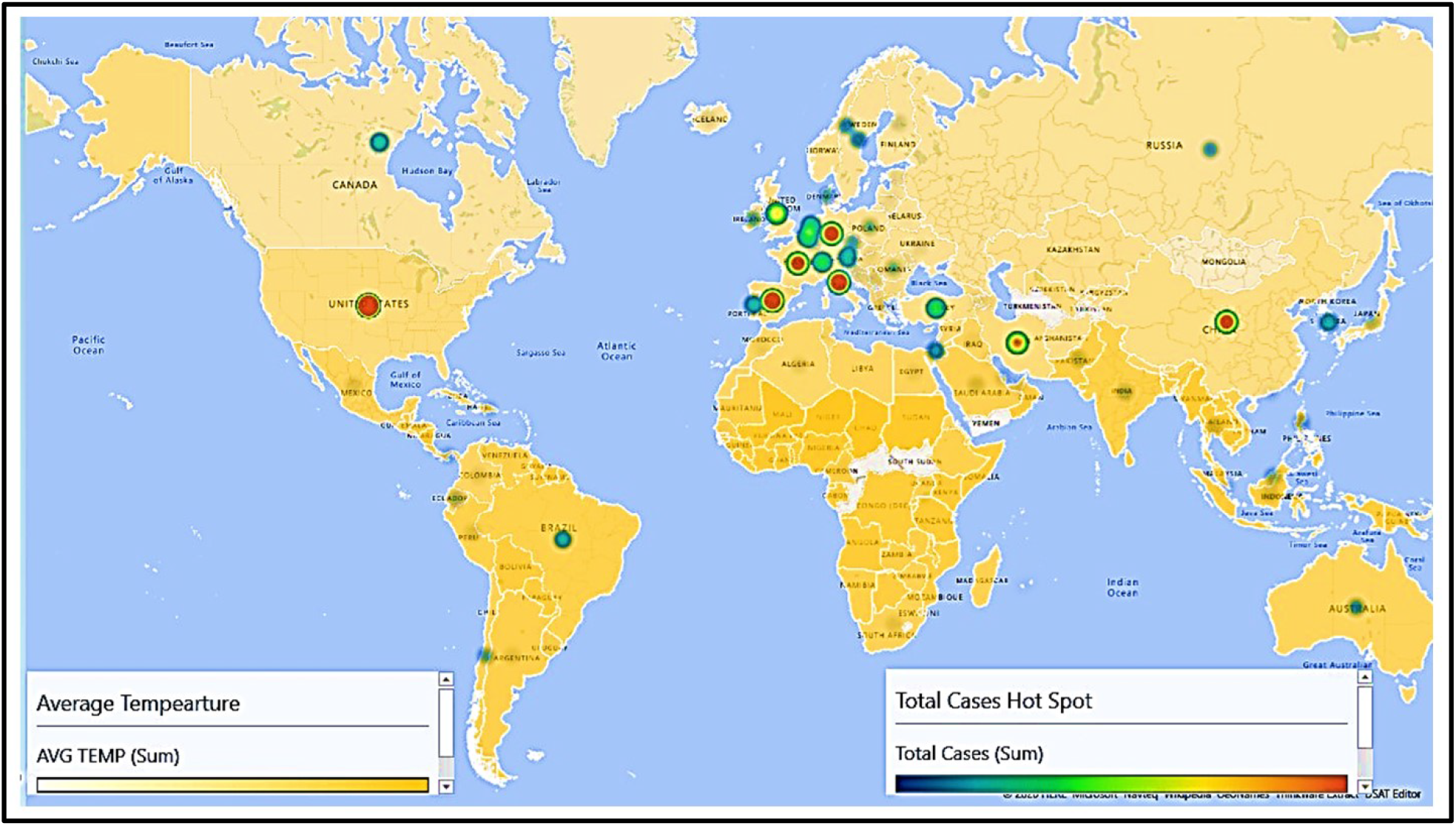
World Heat Map representing the relation between total cases and temperature

### Understanding the relation between total recovery, death and critical cases with average temperature

With **Figure 7, Figure 8, Figure 9** total number of recovery, death, and critical COVID-19 cases are presented respectively and a non-linear fit regression model has been done to understand the relation between them. The *P*-value for the plot Recovery vs Temperature analysis was lower than 0.001 which is statistically significant stating the dependency of recovery rate with that of the temperature. The residual plots also indicate normally distributed data with significance in versus fit. But the R^2^ value found was not satisfactory. Indeed recovery rate depends on various factors like immunity, medical facilities, medical Infrastructure, and therapeutic strategies. Total recovery from COVID-19 infection was found to be different with different average temperatures. Most recovery was registered for temperature between 60 °F to 70 °F. Death and critically affected COVID-19 cases could also be related as shown in **Figure 8 & Figure 9**. But Death and Critical cases can’t be directly correlated with that of the temperature as the P-value for the plot is greater than .05, hence in-significant.

**Figure 7:**
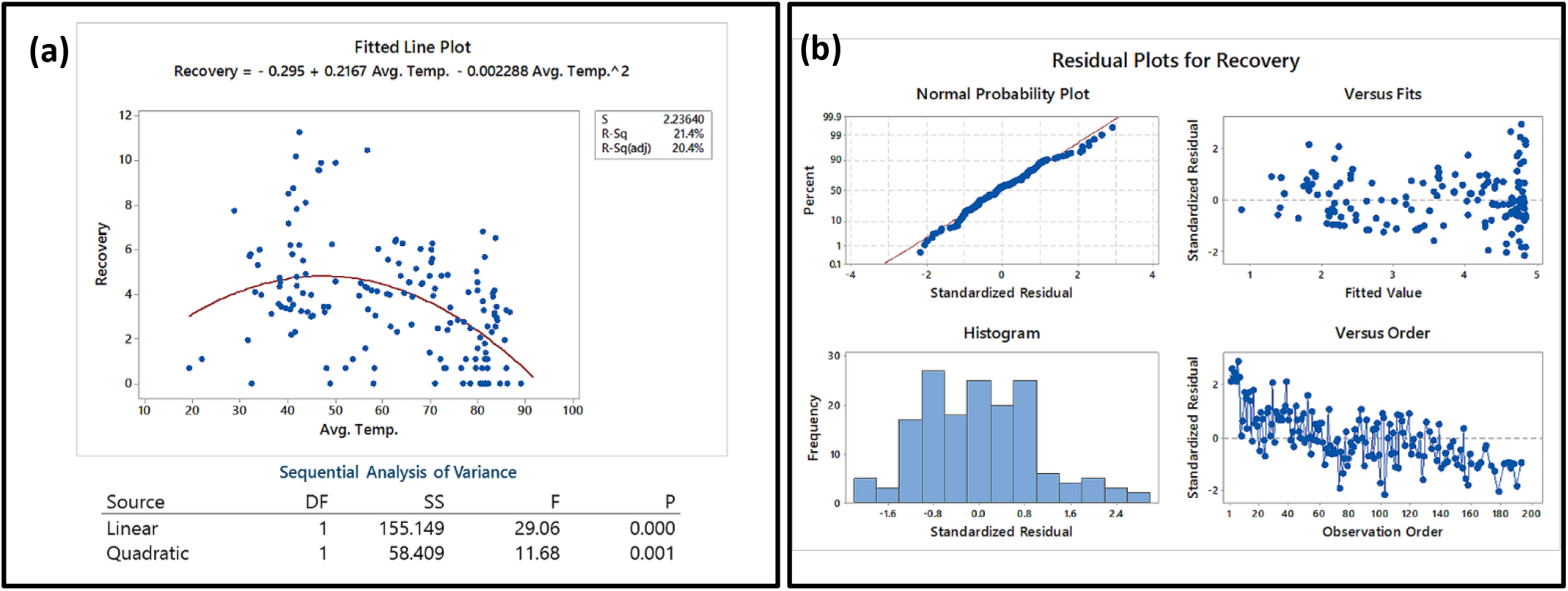
**(a)** The relationship between average temperature and Recovery of COVID-19 **(b)** The residual plots for the statistical analysis

**Figure 8:**
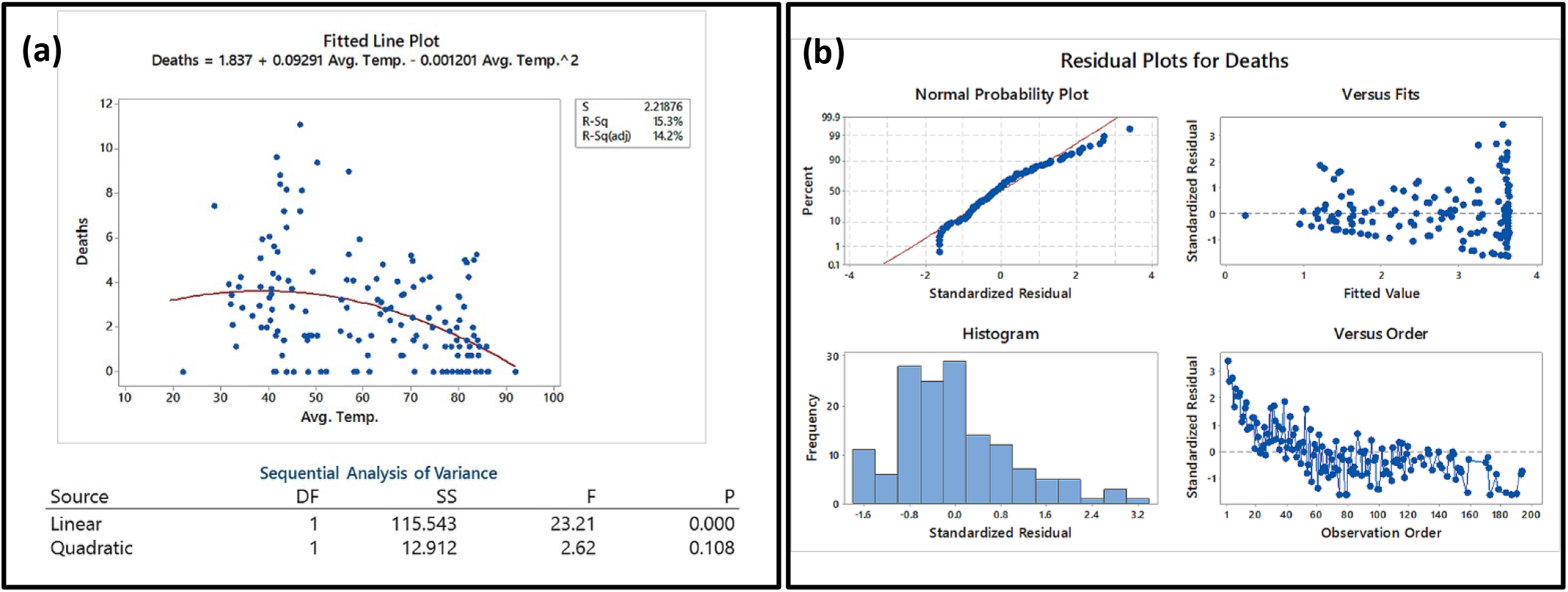
The relationship between average temperature and Death of COVID-19 transmission in the world **(b)** The residual plots for the statistical analysis

**Figure 9:**
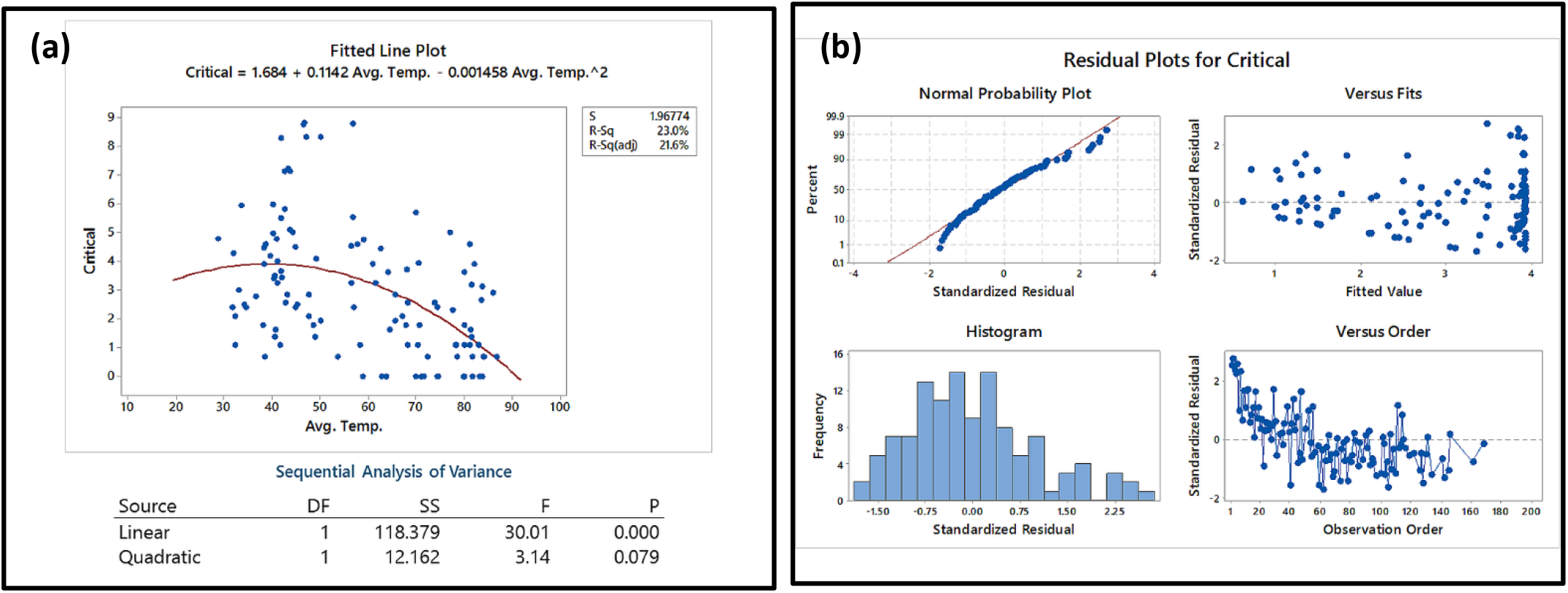
The relationship between average temperature and Critical Cases of COVID-19 transmission in the world **(b)** The residual plots for the statistical analysis

Although, these observations pointing to the fact that the average temperature of a region might be playing a critical role in fast recovery of COVID-19 affected patients, other parameters like poor medical facilities, lack of immunity, poor sanitation and should also be considered. Also, part of the critically affected COVID-19 patients were suffering from other complications that lower their immunity towards the SARS-CoV-2 virus.

### Humidity and total case

The effect of humidity on COVID-19 infection was also noticed and the data were plotted with regression and presented with **Figure 10**. The scatter plot indicated that humidity has less effect on the COVID-19 infection. The p-value was also found to be greater than 0.05 which is statistically insignificant. The residual plot also indicates an unskewed histogram and a non-linear normal probability plot. This observation indicated that environmental parameter like humidity has less effect on the spreading of COVID-19 infection.

**Figure 10:**
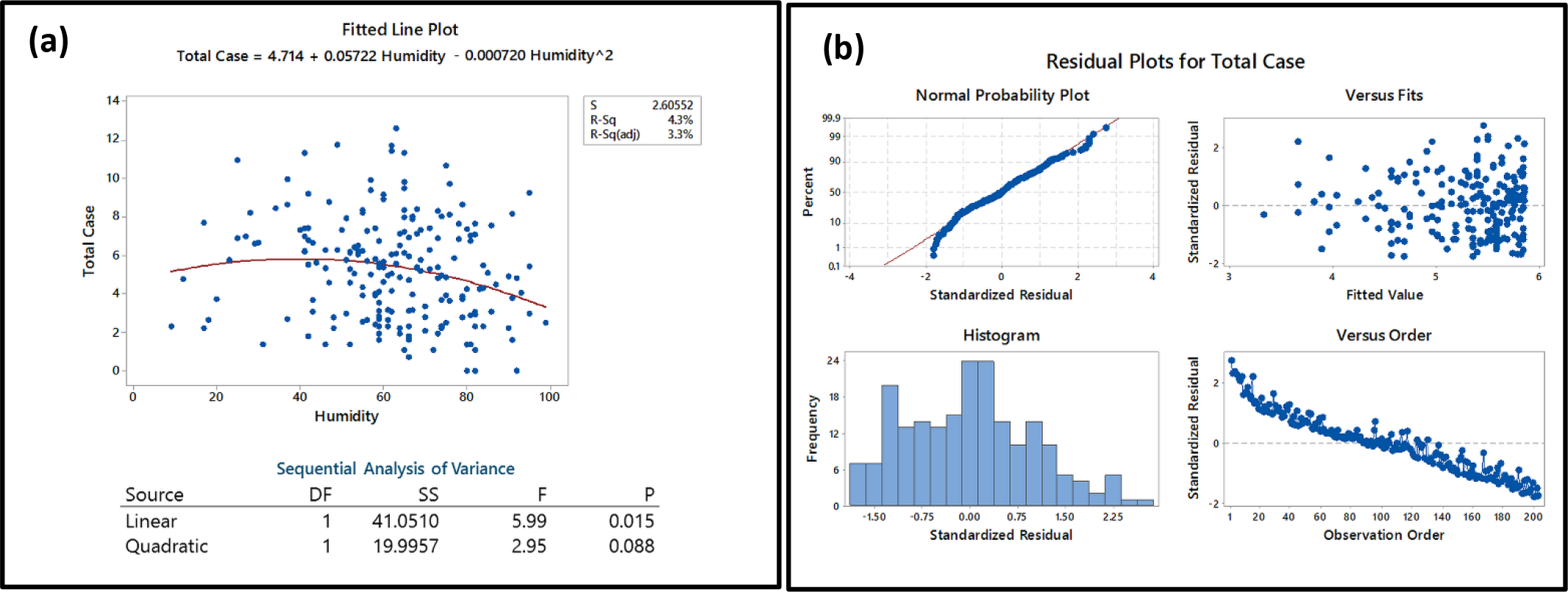
The relationship between average Humidity and Total Cases of COVID-19 transmission in the world **(b)** The residual plots for the statistical analysis

## Conclusion

The study can conclude that temperature has a noteworthy influence on the transmission of COVID-19. Not only transmission, but temperature also plays a significant role in the recovery rate, deaths, and critical cases all around the Globe. The relation is indeed a non-linear indicating that there is a typical favorable temperature that might be contributing to the transmission. Results do suggest that regions having low temperatures are more prone to infection than that of the regions with higher temperatures. Although several other factors like hospital facilities, Government awareness, Medical facilities should also be considered along with the temperature. Apart from this immunity is also playing a major factor in transmission. The population with higher immunity is less affected. Thus we can say that the emergence of the outbreak throughout the world may be narrowly related to the respective local temperature but other important factors are also playing a crucial role in the spread and control of the disease. Temperature only affects the rate of spread and recovery. We can conclude that countries and regions with a lower temperature must take more serious steps and control measures to prevent this pandemic as there specific temperature may be favorable for the spread of the virus.

## Data Availability

All Data are available.

https://www.worldometers.info/coronavirus/

## Acknowledgment

We would like to acknowledge the “**Centre of Excellence in Phase Transformation and Product Characterization, TEQIP-III”, Jadavpur University**, Kolkata, for providing fellowship and financial support during this work. We would also like to acknowledge every lab member of Biomaterial, Cell Culture, and Microbiology lab of the School of Bioscience and Engineering, Jadavpur. We would like to acknowledge Scientist Manish C Bagchi for his help during the work. All the authors would like to thank all the medical workers and emergency service providers around the globe for their incomparable contribution towards the society and mankind while combating COVID-19.

## Reference

1. Malik YS, Sircar S, Bhat S, Vinodhkumar OR, Tiwari R, Sah R, et al. Emerging Coronavirus Disease (COVID-19), a pandemic public health emergency with animal linkages: Current status update. 2020;

2. McMichael AJ, Woodruff RE. Climate change and infectious diseases. In: The social ecology of infectious diseases. Elsevier; 2008. p. 378–407.

3. McMichael AJ, Woodruff RE, Hales S. Climate change and human health: present and future risks. Lancet. 2006;367(9513):859–69.

4. Portier CJ, Tart KT, Carter SR, Dilworth CH, Grambsch AE, Gohlke J, et al. A human health perspective on climate change: a report outlining the research needs on the human health effects of climate change. J Curr Issues Glob. 2013;6(4):621.

5. La V-P, Pham T-H, Ho M-T, Nguyen M-H, P Nguyen K-L, Vuong T-T, et al. Policy Response, Social Media and Science Journalism for the Sustainability of the Public Health System Amid the COVID-19 Outbreak: The Vietnam Lessons. Sustainability. 2020;12(7):2931.

6. Jebril N. World Health Organization declared a pandemic public health menace: A systematic review of the coronavirus disease 2019 “COVID-19”, up to 26th March 2020. Available SSRN 3566298. 2020;

7. Wang M, Jiang A, Gong L, Luo L, Guo W, Li C, et al. Temperature significant change COVID-19 Transmission in 429 cities. medRxiv. 2020;

8. Tan J, Mu L, Huang J, Yu S, Chen B, Yin J. An initial investigation of the association between the SARS outbreak and weather: with the view of the environmental temperature and its variation. J Epidemiol Community Heal. 2005;59(3):186–92.

9. Zhang L, Lin D, Sun X, Curth U, Drosten C, Sauerhering L, et al. Crystal structure of SARS-CoV-2 main protease provides a basis for design of improved $α$-ketoamide inhibitors. Science (80-). 2020;368(6489):409–12.

10. Worldometer. In 2020. p. https://www.worldometers.info/coronavirus/. Available from: https://www.worldometers.info/coronavirus/

11. List of cities by average temperature [Internet]. Available from: https://en.wikipedia.org/wiki/List_of_cities_by_average_temperature

12. NOAA’s National Centers for Environmental Information (NCEI) [Internet]. Available from: https://www.ncdc.noaa.gov/

